# Stroke survivors experience elevated levels of loneliness: a multi-year analysis of the National Survey for Wales

**DOI:** 10.1101/2020.07.02.20145417

**Authors:** Christopher Byrne, Christopher W. N. Saville, Rudi Coetzer, Richard Ramsey

**Affiliations:** Wales Institute for Cognitive Neuroscience, School of Psychology, Bangor University, Bangor, Gwynedd, Wales, LL57 2AS, United Kingdom; North Wales Brain Injury Service, Colwyn Bay Hospital, Colwyn Bay, Conwy, LL29 8AY; North Wales Clinical Psychology Programme, Bangor University, Bangor, Gwynedd, Wales, LL57 2AS, United Kingdom; Department of Psychology, Macquarie University, Sydney, NSW 2109, Australia

**Keywords:** loneliness, stroke, acquired brain injury, National Survey for Wales

## Abstract

Despite clinical observation that stroke survivors frequently experience loneliness, there is no large-scale empirical evidence to support this observation. To address this issue, we completed two pre-registered analyses of a nationally representative annual survey that included a self-report measure of loneliness (N>21000). Across two separate cohorts, the results consistently showed that human stroke survivors report higher levels of loneliness compared to healthy individuals, and this relationship could not be accounted for by demographic factors (e.g., age, sex) or objective measures of social isolation (e.g., marital status, number of household members). These findings demonstrate that elevated levels of loneliness post-stroke are robust in that they replicate in large nationally representative samples and cannot be reduced to objective measures of social isolation. The work has clinical and societal relevance by suggesting that loneliness post-stroke is unlikely to be adequately “treated” if only the quantity and not the quality of social experiences are considered.

## Introduction

Relationships with other people – social connections – are crucial to health and happiness. Indeed, a perceived lack of social connection (loneliness) is a risk factor for cognitive decline and dementia and is associated with increased morbidity and mortality (Holt-Lunstad et al., 2010; 2015). Although many studies have investigated the prevalence of loneliness in older adults and those with dementia, it remains unclear how widespread loneliness symptoms are across a range of illnesses, including other types of neurological condition. For instance, despite clinical observation that stroke survivors experience many symptoms of loneliness, to date there is no large-scale empirical evidence to support this observation. To address this issue, the current study analysed data from two successive years of the National Survey for Wales (NSW; N>21000), in order to estimate the extent to which stroke survivors experience loneliness more than healthy individuals, how it compares to other common illnesses and whether it can be accounted for by demographic factors such as age, sex and environmental factors such as population density, size of household and internet access. The work impacts basic research by informing the neural and cognitive determinants of loneliness and it also has clinical and societal relevance by providing the first comprehensive estimates of how the perceptions of social connections are shaped by stroke.

The terms social isolation and loneliness are often used interchangeably. Although conceptually related, loneliness and social isolation are dissociable (Perissinotto & Covinsky, 2014). Social isolation is objectively quantifiable and relates to factors such as the quantity and frequency of social contact as well as proximity to others. By contrast, loneliness reflects a person’s evaluation of their social relationships in a way that identifies that their social needs are not being met satisfactorily (Cacioppo & Patrick, 2008; Valtorta, Kanaan, Gilbody, & Hanratty 2016). Loneliness, therefore, reflects the interpretation of an individual’s social surroundings as being inadequate to their needs. In other words, measures of loneliness index the perceived quality, rather than quantity, of social connections.

The relationship between social isolation and loneliness is not straightforward. On average, higher loneliness is reported in those who live alone. However, many who live alone do not report elevated levels of loneliness and many who live with others report feeling lonely (Perissinotto et al., 2012). Moreover, although transient loneliness may be adaptive under some conditions because it triggers attempts to reaffiliate with others (Qualter et al., 2015), studies have demonstrated that prolonged loneliness can result in serious health outcomes that are not explained by objective measures of social isolation. For example, studies have shown that loneliness predicts physiological responses associated with stress (e.g., elevated blood pressure, rise in cortisol), as well as maladaptive cognition and behaviour (e.g., perception of social interactions as unpleasant, reduced physical activity and lifetime satisfaction) above and beyond objective measures of social isolation (Bruine de Bruin, Parker, & Strough, 2020; Cacioppo, Cacioppo, Capitanio, & Cole, 2015; Cacioppo & Hawkley, 2009). Further, loneliness has been shown to be associated with increased risk of morbidity and mortality (Hienrich & Gullone, 2006; Holt-Lunstad et al., 2015). Therefore, to understand the relationship between one’s social environment and other factors such as physiological responses, cognitive and brain mechanisms or health outcomes, it is not sufficient to simply quantify social contact. Instead, it is also important to capture how an individual interprets their social environment (Bruine de Bruin, Parker, & Strough, 2020; Cacioppo & Patrick, 2008; Hawkley et al., 2008). Consequently, the distinction between social isolation and loneliness is a valuable one to make when attempting to make progress understanding basic biological systems as well as developing clinical interventions.

Given the negative consequences that loneliness has for the health and wellbeing of society, research has recently started to uncover the psychological and biological mechanisms that control the experience of loneliness, as well as the relationship between loneliness and other disease states. One line of research, for example, has started to establish the biological mechanisms that underpin the exacerbation of disease in reaction to loneliness. Three major biological systems that are sensitive to prolonged loneliness; the neuroendocrine (HAP) axis, the immune system, and the autonomic nervous system (Cacioppo, Cacioppo, Capitanio, & Cole, 2015; Friedler, Crasper & McCullough, 2015). Furthermore, not only does loneliness demonstrate a causal relationship with disease, it also appears to mediate the trajectory of recovery and deterioration from such diseases (Friedler, Crasper & McCullough, 2015). As such, the experience of loneliness provides a significant stress to the body, which is comparable to other risk factors for poor health such as obesity and smoking (Cacioppo, Grippo, London, Goossens & Cacioppo, 2015). Importantly, because loneliness reflects a perception of one’s social environment, it cannot be “treated” by simply reshaping the quantity of social contact. Instead, to produce more effective clinical interventions, we must understand the cognitive and brain mechanisms that underpin the perception of loneliness.

A common approach to understanding brain function involves comparing behavioral and neural profiles between healthy individuals and those in a diseased or clinical condition. To date, research that has linked the experience of loneliness with neurological conditions has focused on individuals with dementia who are typically aged 65 and over. Reports from the Alzheimer’s Society indicate that 38% of those with dementia experience high levels loneliness (Kane & Cook, 2013). Current evidence indicates a complex connection between loneliness and dementia. Grande and colleagues (2018) demonstrated that, after controlling for age, sex, years of education and comorbidities, those with early mild cognitive impairment (MCI) and who lived alone, had a 50% increased risk of developing dementia, when compared to those who lived with someone. However, the relationship between dementia and loneliness may be bidirectional. It may be that loneliness is a prodromal presentation in the early stages of the development of dementia. Therefore, examining loneliness in other neurological conditions with different etiologies may prove useful when exploring the relationship between the loneliness and brain pathology. Given the limited number of neurological conditions that have been studied to date in relation to loneliness, the extent to which loneliness is a common experience across different types of brain related illnesses, such as stroke, remains unclear.

Stroke, ischemic and haemorrhagic, is a leading cause of death and disability in the United Kingdom (UK). Incidence rates of stroke have been calculated at over 100,000 of the UK population each year (Stroke Association, 2018). Whilst predominately associated with the older population, Public Health England (PHE) found that over a third of strokes occur in middle-aged adults (40 to 69 years old) (PHE, 2018). Thankfully, with the advancement of medical interventions, more people are now surviving stroke. Although the incidence rate of stroke is high, deaths have declined by 49% over the last 15 years with over 1.2 million stroke survivors in the UK (Stroke Association, 2018). However, many stroke survivors are left with the physical, cognitive and emotional sequelae.

The functional consequences of stroke are heterogenous and determined by a complex array of pre- and post-stroke factors such as etiology, brain region affected, social situation and level of pre-morbid functioning, to name a few. Typically, improvements in functional status from the acute stage of recovery to a 3-year follow-up are demonstrated. However, latent psychosocial difficulties such as loneliness and depression have been reported to arise in the later stages of recovery (Harrick et al, 1994). For example, in a longitudinal study of 21 individuals following a severe brain injury, Harrick and colleagues (1994) showed that loneliness and depression increased over time to become the most frequently reported problems at three-year follow-up. Such findings provide suggestive evidence for a relationship between brain injury and loneliness, but the evidence remains limited by a relatively modest sample size.

Other methodological approaches have also provided suggestive evidence for a link between loneliness and stroke by estimating the extent to which social isolation is a risk factor for stroke. In a meta-analysis of longitudinal observational studies, social isolation – that is, a reduced quantity of social contact – was shown to be a risk factor for stroke (Valtorta, Kanaan, Gilbody, Ronzi, & Hanratty, 2016). Given the links between social isolation and loneliness, these data are also suggestive of a link between loneliness and stroke, but direct and comprehensive evidence is still lacking. As such, although there is suggestive evidence for a link between stroke and loneliness to date, the gaps in current understanding emphasise the clinical need to study loneliness comprehensively in the stroke population, in order to estimate the determinants of loneliness and the scale of the difficulties experienced.

The overarching aim of this work was to provide the first large-scale and comprehensive estimate of loneliness in stroke survivors. To circumvent the practical difficulties associated with recruiting a sufficient number of stoke survivors and appropriate control participants to provide powerful inferences, the current study uses nationally representative survey data (the National Survey for Wales; NSW). The NSW is an annual survey of approximately 10,000 individuals who live in Wales, that spans a range of sociodemographic and health questions. Of particular relevance for the purpose of the present work is that a loneliness questionnaire was included in two separate years of the survey (Gierveld & Tilburg, 2006). Our general approach, which we pre-registered online before beginning the analysis, was to use one year’s data to test our initial predictions and then use the subsequent year’s data to perform a confirmatory analysis. Given the low levels of reproducibility in psychology and neuroscience and the common use questionable research practices (Munafo et al., 2017; Open Science Collaboration, 2015; Simmons et al., 2011), the advantage of taking this multi-cohort methodological approach is that replication is built into our design, which considerably strengthens the inferences that we are able to make (Zwaan et al., 2018).

We used these survey data to address two related aims. First, we wanted to estimate the degree of loneliness experienced in stroke survivors and compare it to a healthy population, as well as a different ill-health condition such as arthritis, which does not have a neurological cause. We hypothesized a graded set of differences between these groups such that loneliness would be higher following stroke than arthritis, but both would be higher than healthy individuals. Second, to further qualify the experience of loneliness in stroke survivors, we wanted to estimate the extent to which loneliness following stroke could be explained by demographic factors such as sex, age, as well as proxy measures for the quantity of social isolation (e.g., number of people in a household, population density and access to the internet). Stronger evidence for a relationship between stroke and loneliness per se, rather than a common correlate, would be provided if the relationship is not explained by other demographic or social isolation factors.

Establishing a clear link between brain injuries like stroke and loneliness informs basic research as well as treatments in clinical settings. In terms of basic understanding, it is important to know the range of antecedents – i.e., different forms of brain injury – that may relate to loneliness, as it may shed some light on what mechanisms give rise to the experience social connectedness or lack of it. The clinical importance is that should a clear link exist between stroke and loneliness, then a clinician’s job should be to take seriously the possible social consequences of a stroke, as ignoring them are likely to have serious health implications. Further, it may bolster support for the idea that a clinician’s toolkit may require different clinical solutions e.g., social prescriptions, in addition to standard treatment, as well as how such prescriptions should be structured e.g., with a focus on developing a greater quality, rather than quantity, of social connections.

## Method

### Overview of the general methodological approach

The methodological approach involved secondary data analyses from two existing data sets collected by the NSW, Office of National Statistics (ONS). The NSW is designed to provide a standardized, nationally representative sample of the entire adult population (aged 16 and over) of Wales. The design of the NSW is described in greater detail in the Welsh Government Technical Report by Helme & Brown (2018). In order to access the data sets, an account was registered with the UK Data Service (https://ukdataservice.ac.uk), where the data sets can be downloaded online. Ethical approval for the study was granted by the Bangor University ethics review board.

A two-phase approach was adopted that involved combining exploratory (Study 1) and confirmatory (Study 2) phases. Both studies were pre-registered in advance of running the analyses to enable hypotheses, study design and analysis choices to be defined in advance. The aim of pre-registration is to make a formal and explicit division between exploratory and confirmatory analyses, as well as to avoid HARKing (Hypothesising After Results are Known). The exploratory phase (Study 1) used the NSW 2016/17 data and was pre-registered on the 14^th^ February 2019 (https://aspredicted.org/jt479.pdf). The confirmatory phase (Study 2) used the NSW 2017/18 data and was pre-registered on 28^th^ October 2019 (https://aspredicted.org/xq5bd.pdf). Even though it might seem unnecessary to pre-register our analyses for the exploratory phase, we felt that it was worthwhile for two reasons. First, even prior to Study 1, we already had quite well-formed hypotheses regarding the relationship between loneliness and stroke, which we felt should be recognised in advance of running the analyses. Second, given the NSW datasets are very large and span approximately 2000 variables, we felt that it was worth registering in advance how the vast majority of variables were not of interest to our primary questions.

### Open data and analyses

The data that this study uses are of open and freely available for download via the UK Data Service website (https://ukdataservice.ac.uk). Therefore, if other researchers wish to pursue alternative research questions and analyses, then they are able to do so. In addition, following recent open science recommendations (Munafo et al., 2017), we provide an R Markdown file that documents each step of our analytical pipeline in order to aid transparency, sharing of data and to allow others to reproduce any aspect of our analyses that they want (https://osf.io/69kcd/).

### Characteristics of the samples and primary groups of interest

We were interested in comparing levels of reported loneliness in stroke survivors to several comparison groups. Primary comparison groups included individuals with no longstanding illnesses, those with a non-brain-based illness (arthritis), and a combined average of all individuals reporting any longstanding illness. In this section, we summarise the characteristics of these groups across both datasets. In total, there were 21,874 participants across both NSW datasets (2016/17 & 2017/18). The 2016/17 NWS dataset (Study 1) consisted of 10,493 study participants. There were 4878 who reported no longstanding illness and 5550 who report some form of previous illness. Sixty-five study participants did not answer the question relating to health status (4 not asked due to routing of question, 38 reported that they did not know, 20 terminated the interview early and 3 refused to answer the question). Of those that reported a previous illness, 110 participants reported a history of stroke, of which 79 answered questions relating to levels of loneliness from the loneliness questionnaire. A total of 1298 participants reported having arthritis, of which 1094 answered the loneliness questionnaire. The NSW 2017/18 dataset (Study 2) consisted of 11,381 study participants. There were 5347 who reported no longstanding illness and 5971 that report a previous longstanding illness. Sixty-three study participants did not answer questions relating to health status (4 not asked due to routing of question, 30 terminated the interview early, 25 reported they did not know and 4 refused to answer the question). There were 134 participant who reported a history of stroke, of which 117 answered question from the loneliness questionnaire. A total of 1408 participants reported having Arthritis, of which 1267 answered the loneliness questionnaire. A complete set of demographic information for each dataset separated for each condition (stroke, arthritis and no longstanding illness) is reported in *Table 1*.

**Table 1.** Demographic information for Stroke, Arthritis and No Longstanding Illness group.

|  | Stroke |  | Arthritis |  | No longstanding illness |  |
| --- | --- | --- | --- | --- | --- | --- |
|  | 2016/17 | 2017/18 | 2016/17 | 2017/18 | 2016/17 | 2017/18 |
| **Gender** |  |  |  |  |  |  |
| Male | 60 (54.5%) | 78 (58.2%) | 412 (31.8%) | 417 (29.6%) | 2182 (44.7%) | 2478 (46.4%) |
| Female | 50 (45.5%) | 56 (41.8%) | 885 (68.2%) | 991 (70.4%) | 2695 (55.3%) | 2868 (53.6%) |
| **Age category** |  |  |  |  |  |  |
| 16to29 | 0 (0%) | 0 (0%) | 7 (0.5%) | 12 (0.9%) | 888 (18.2%) | 935 (17.5%) |
| 30to39 | 2 (1.8%) | 2 (1.5%) | 32 (2.5%) | 32 (2.3%) | 888 (18.2%) | 972 (18.2%) |
| 40to49 | 8 (7.3%) | 8 (6.0%) | 80 (6.2%) | 78 (5.5%) | 906 (18.6%) | 919 (17.2%) |
| 50to59 | 17 (15.5%) | 24 (17.9%) | 203 (15.6%) | 254 (18.0%) | 835 (17.1%) | 959 (17.9%) |
| 60to69 | 27 (24.5%) | 36 (26.9%) | 391 (30.1%) | 370 (26.3%) | 724 (14.8%) | 835 (15.6%) |
| 70to79 | 30 (33%) | 38 (28.4%) | 392 (30.2%) | 405 (28.8%) | 443 (9.1%) | 527 (9.9%) |
| 80plus | 23 (20.9%) | 26 (19.4%) | 193 (40.9%) | 257 (18.3%) | 194 (4%) | 200 (3.7%) |
| **Marital Status** |  |  |  |  |  |  |
| Single | 14 (12.7%) | 21 (15.7%) | 151 (11.6%) | 161 (11.4%) | 1637 (33.6%) | 1794 (33.6%) |
| Married | 43 (39.1%) | 59 (44.0%) | 560 (43.2) | 603 (42.8%) | 4722 (45%) | 2484 (46.5%) |
| Same sex relationship | 0 (0%) | 0 (0.0%) | 2 (0.2%) | 2 (0.1%) | 12 (0.1%) | 4 (0.1%) |
| Separated | 2 (1.8%) | 2 (1.5%) | 32 (2.5%) | 31 (2.2%) | 322 (3.1%) | 117 (2.2%) |
| Divorced | 23 (20.9%) | 22 (16.4%) | 233 (18%) | 252 (17.9%) | 1339 (12.8%) | 565 (10.6%) |
| Widowed | 28 (25.5%) | 30 (22.4%) | 319 (24.6%) | 359 (25.5%) | 1340 (12.8%) | 381 (7.1%) |
| **Level of Deprivation** |  |  |  |  |  |  |
| Most deprived 20% | 20 (18.2%) | 24 (17.9%) | 252 (19.4%) | 311 (22.1%) | 688 (14.1%) | 799 (14.9%) |
| Q2<sup>†</sup> | 18 (16.4%) | 37 (27.6%) | 245 (18.9%) | 305 (21.7%) | 847 (17.4%) | 942 (17.6%) |
| Q3<sup>†</sup> | 31 (28.2%) | 21 (15.7%) | 312 (24%) | 291 (20.7%) | 1116 (22.9%) | 1143 (21.4%) |
| Q4<sup>†</sup> | 20 (18.2%) | 28 (20.9%) | 278 (21.4%) | 265 (18.8%) | 1266 (26%) | 1312 (24.5%) |
| Least deprived 20% | 21 (19.1%) | 24 (17.9%) | 211 (16.3%) | 236 (16.8%) | 961 (19.7%) | 1151 (21.5%) |
| **Population Density** |  |  |  |  |  |  |
| <2000 sparsest context | 16 (14.5%) | 14 (10.4%) | 202 (15.6%) | 173 (12.3%) | 791 (16.2%) | 701 (13.1%) |
| <2000 | 10 (9.1%) | 19 (14.2%) | 120 (9.2%) | 130 (9.2%) | 567 (11.6%) | 565 (10.6%) |
| 2000 to 9,999 | 23 (20.9%) | 25 (18.7%) | 309 (23.8%) | 326 (23.2%) | 1046 (21.4%) | 1120 (20.9%) |
| 10,000 to 24,999 | 16 (14.5%) | 21 (15.7%) | 233 (18%) | 264 (18.8%) | 902 (18.5%) | 948 (17.7%) |
| 25,000 to 99,999 | 24 (21.8%) | 27 (20.1%) | 236 (18.2%) | 251 (17.8%) | 849 (17.4%) | 968 (18.1%) |
| At least 100,000 | 21 (19.1%) | 28 (20.9%) | 198 (15.3%) | 264 (18.8%) | 723 (14.8%) | 1045 (19.5%) |
| **Number of people in household** |  |  |  |  |  |  |
| Sole occupant | 50 (45.5%) | 64 (47.8%) | 585 (45.1%) | 651 (46.2%) | 1097 (22.5%) | 1261 (23.6%) |
| 2 people | 48 (43.6) | 58 (43.3%) | 554 (42.7%) | 599 (42.5%) | 1733 (35.6%) | 1925 (36.0%) |
| 3 to 5 people | 11 (10%) | 12 (9.0%) | 154 (11.9%) | 151 (10.7%) | 1957 (40.2%) | 2040 (38.2%) |
| More than 5 people | 1 (0.9%) | 0 (0.0%) | 3 (0.2%) | 7 (0.5%) | 84 (1.7%) | 121 (2.3%) |
| **Internet access** |  |  |  |  |  |  |
| yes | 71 (65.1%) | 84 (62.7%) | 943 (72.7%) | 1041 (74.0%) | 4391 (90%) | 4889 (91.5%) |
| no | 38 (34.9%) | 50 (37.3%) | 354 (27.3%) | 365 (26.0%) | 487 (10%) | 457 (8.5%) |

† Level of Deprivation Q2 – 2nd Quintile; Q3 – 3rd Quintile; Q4 – 4th Quintile.

### Measures

#### Loneliness

The dependent/outcome variable, loneliness, was measured using the 6-item De Jong Gierveld Loneliness Scale (DJGLS), which is a shortened version of the 11-item DJGLS (Gierveld & Tilburg, 2006). The 6-Item scale consists of statements relating to one’s interpretation of their social situation. For example, “*I miss having people around me*” and “*there are enough people I feel close to*”. Each item is answered categorically (Yes / More or less / No). Answers of ‘yes’ or ‘more or less’ are given a score of 1. Answers of ‘no’ are scored as 0. The total score is summed to provide a total loneliness score. Therefore, the total score can range from 0 – 6, with higher scores reflecting higher levels of reported loneliness. The 6-item DJGLS’ validity and reliability has been empirically tested using data from two large scale surveys (N= 9,448). The 6-item DJGLS was found to be a reliable and valid measurement instrument to examine the subjective evaluation of individuals’ social situation (loneliness), with reliability α coefficients between .70 and .76 (Gierveld & Tilburg, 2006).

#### Health status

Health status was measured through an open-ended item of the NSW questionnaire: *“What* (*other*) *health problem or disability do you have?”*. The health condition was recorded, and a derived variable was subsequently created logging the presence, or indeed absence, of health conditions for each participant. The current study specifically identified three health conditions of interest: ‘stroke’, ‘arthritis’ and ‘no longstanding illness’. Arthritis was selected as a comparison ill-health group as the pathological mechanism of the condition primarily affects the synovial joints and does not directly involve the central nervous system. In addition, similar to stroke, arthritis affects a similar demographic population in that it is highly associated with aging, which is further demonstrated in *Table 1*.

#### Demographic factors

The current study also included a range of demographic factors in our analyses, such as ‘gender’ (male and female), ‘age’, ‘marital status’ and ‘level of deprivation of neighborhood’ (Welsh Index of Multiple Deprivation; WIMD). Age was included as a continuous variable. Level of deprivation (WIMD) was categorised into five quintiles ranging from: quintile 1 (most deprived 20%) to quintile 5 (least deprived 20%). Marital status was also categorised as follows; ‘Single’, ‘Married’, and ‘Ended’. Single refers to not being in a relationship, married includes those who are married or in a same-sex civil partnership and ended includes those that are separated, divorced or widowed.

#### Social isolation

In addition to relevant sociodemographic variables, factors relating to social isolation (e.g., the objective quantity and frequency of social connections) were also included in our analyses. We used several proxy measures of social isolation Iing ‘number of persons in household’, ‘population density of area’ and ‘access to internet connection’. The number of people in the household was included as a continuous variable. Population density was also categorized as; ‘less than 2000’, ‘2000 to 9,999’, ‘10,000 to 24,999’, ‘25,000 to 99,999’ and ‘over 100,000’. Access to internet was coded in a binary fashion based on the presence or absence of a connection.

### Analytical approach

Following proposals by Gigerenzer (2018), we avoid interpreting results based solely on p values that result from hypothesis tests and a binary distinction between “significant” and “non-significant”. Instead, we base the direction and strength of our interpretation on a range of metrics, which include a mixture of descriptive and inferential statistics. We consider hypothesis tests and p values in combination with an estimation approach, which emphasises the importance of estimating effect sizes (in original and standardised units) along with a measure of precision using 95% confidence intervals (Cumming, 2012; 2014; McElreath, 2016; Kruschke & Liddell, 2018). Further, we have embedded replication into the design of the study by using two independent cohorts (Zwaan et al., 2017) and we meta-analyse the primary effects of interest across cohorts (Cumming, 2012). Replication and meta-analysis further aid the confidence that we can have in our findings and the credibility of the conclusions by increasing the robustness of our estimated effects.

Descriptive analyses were performed to describe the characteristics of the sample including age, gender, marital status and social isolation (see Table 1). We also compared loneliness scores as a function of demographic variables in the stroke and non-stroke populations. To do so, we calculated mean differences in loneliness scores as a function of demographic variables, and we ran several Chi Square tests. This latter analysis has the potential to identify demographic groups who may be more vulnerable to loneliness in general and following stroke. Of course, given that the total number of post-stoke individuals in each cohort of data is approximately100, any sub-analysis of the stroke population is only going to provide suggestive, rather than compelling evidence. Across both studies, there was no consistent pattern of results that emerged regarding loneliness scores in the stroke population according to demographic factors (Supplementary Tables 1 and 2).

**Table 2:** Loneliness scores across years and health conditions.

| *study* | *year* | *group* | *n* | *mean* | *sd* | *se* | *ci* |
| --- | --- | --- | --- | --- | --- | --- | --- |
| Study1 | 16_17 | no longstanding illness | 4390 | 1.61 | 1.59 | 0.02 | 0.05 |
| Study1 | 16_17 | ill<sup>a</sup> | 4701 | 2.04 | 1.79 | 0.03 | 0.05 |
| Study1 | 16_17 | arthritis | 1094 | 1.98 | 1.80 | 0.05 | 0.11 |
| Study1 | 16_17 | stroke | 79 | 2.33 | 1.84 | 0.21 | 0.41 |
| Study2 | 17_18 | no longstanding illness | 4930 | 1.53 | 1.55 | 0.02 | 0.04 |
| Study2 | 17_18 | ill<sup>†</sup> | 5411 | 1.98 | 1.82 | 0.02 | 0.05 |
| Study2 | 17_18 | arthritis | 1267 | 2.04 | 1.84 | 0.05 | 0.10 |
| Study2 | 17_18 | stroke | 117 | 1.96 | 1.71 | 0.16 | 0.31 |

<sup>†</sup> ill –any longstanding illness. n = number of individuals per group. sd = standard deviation. se = standard error. ci = 95% confidence interval.

To examine the first research question, which was centered on the hypothesis that those with a history of stroke will report higher levels of loneliness when compared to those with arthritis and those with no longstanding illness, a series of independent samples t-tests were completed. We expected loneliness to be higher in stroke than arthritis and both ill-health groups to report greater loneliness than the no longstanding illness group. Therefore, we had directional predictions for all of our paired contrasts and use one-sided t-tests and 95% confidence intervals.

To examine the second research question, which was focused on estimating if stroke made a unique contribution to loneliness above and beyond demographic and social isolation factors, we used hierarchical binary logistic regression analysis. By doing so, we could determine the Odds Ratio (OR) and 95% Confidence intervals (95% CI) between the predictor/independent variables and loneliness scores. Loneliness scores were dichotomized into “not lonely” or “lonely” in order to fit the binary regression model. Participants with DJGLS scores of ≤ 2 were categorised as “not lonely”. Conversely, DJGLS scores of ≥ 3 were categorised as “lonely”. Demographic variables were entered at the first stage of the model (Model 1), followed by social isolation variables (Model 2), then finally presence of health condition (Model 3). Constructing several successive models in this manner made it possible to estimate the influence that stroke has on loneliness scores in addition to demographic and social isolation variables.

Finally, a meta-analysis was completed to synthesize the independent t-test findings from both the exploratory and confirmatory phases in order to provide a weighted estimate of three relevant effect sizes across both datasets. We focused on three group differences (stroke > no longstanding illness; stroke > arthritis, and; stroke > any longstanding illness). To perform the meta-analysis, we used the ‘metafor’ package within the R programming language (Viechtbauer, 2010). We first used the ‘escalc’ function to calculate the mean difference between the groups in each study. Subsequently, we used the ‘rma’ function to fit a random effects model using the default method, which is the restricted maximum-likelihood estimator.

## Results

### General incidence of stroke across both datasets

In total, across both phases, 244 (1.1%) out of the 21,874 participants reported a history of stroke. This figure is in line with incidence rates of stroke in the general UK adult population. The Stroke Association UK report 1.2 million people have a history of stroke in a UK population of 66.4mil (1.8%). Visual analysis of the sample characteristics (see Table 1) revealed little difference in demographic factors between the stroke group and the arthritis group. However, the arthritis group had a higher percentage of females. This is to be expected as the epidemiological studies have demonstrated the sex ratio is typical 3:1 with females more likely affected (van Vollenhoven, 2009). In addition, the no longstanding illness group demographic age was younger when compared to the stroke and arthritis group.

### Exploratory phase (Study 1)

#### Describing loneliness across our groups of interest

The total prevalence of loneliness reported, as defined by scores of 3 or over on the DJGLS, was 44% in the stroke group, 34% in the arthritis group and 27% in the no longstanding illness group. In addition, reported loneliness ratings across the three groups were as follows: stroke (n = 79, x = 2.33, 95%CI [1.92, 2.74]); arthritis (n = 1298, x = 1.98, 95%CI [1.89, 2.08]), and; no longstanding illness (n = 4878, x = 1.61, 95%CI [1.56, 1.66] (*Table 2; Figure 1*).

**Figure 1.**
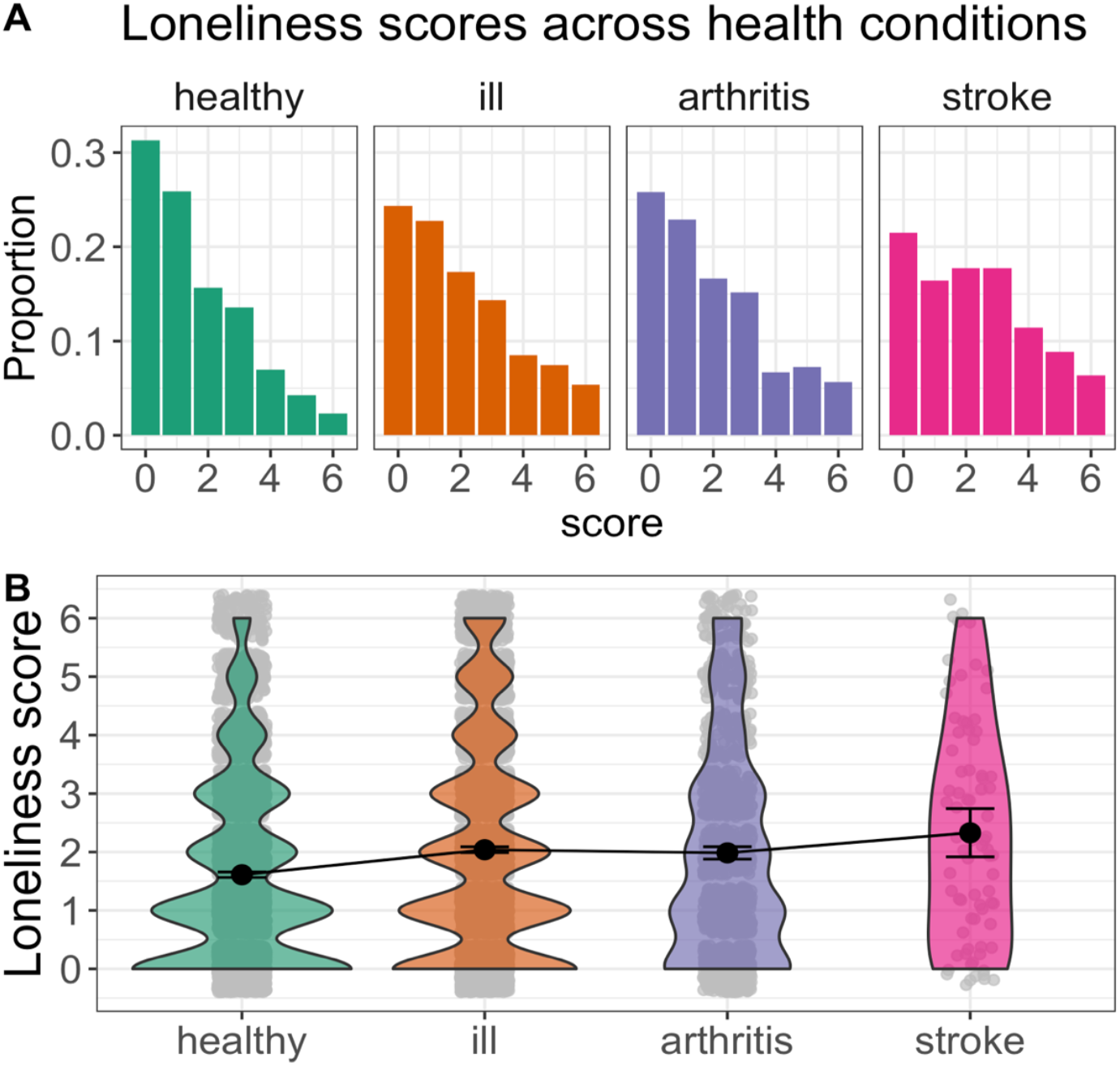
Loneliness scores across health conditions in Study 1. Histograms (A) and violin plots (B) of loneliness scores in Study 1. A) Histogram representing the proportion of reported loneliness score for each health condition. B) Violin plot representing the mean and 95%CI for each health control group in addition to the distribution of scores. healthy = no longstanding illness, ill = any longstanding illness.

### Analysis of research question 1: To what extent do loneliness ratings differ between health conditions

To estimate the size of any differences in loneliness between groups, we ran several inferential statistical analyses. Three independent samples t-tests were performed to estimate pair-wise group differences of interest. First, we compared loneliness scores between the stroke and no longstanding illness groups. The stroke group reporting higher loneliness than the no longstanding illness group t(80.10) = 3.44, p < 0.001, difference in rating = 0.72, 95%CI[0.37, Inf], demonstrating a medium effect with no overlap between groups, *d* = 43, [0.23, 0.67].

Second, we compared the stroke group to the arthritis group. The stroke group showed a difference in loneliness in the expected direction, t(89.06) = 1.61, p = 0.06 difference in rating = 0.34, [-0.01, Inf], demonstrating a small effect with overlap between groups, d=0.19, [-0.04, 0.42].

Third, we compared the arthritis group to the no longstanding illness group. The arthritis group also showed a difference in loneliness rating in the expected direction, t(1545.78) = 6.29, [-0.06, Inf], difference in rating = 0.37, demonstrating a small effect with no overlap between groups, d = -0.23, [0.16, 0.30].

In Study 1, we also ran additional between-group analyses that we did not pre-register. The main focus of these additional analyses was to help contextualise our primary analyses by comparing loneliness scores to a wider set of ill-health conditions. More specifically, we decided to compare loneliness ratings in the stroke group to an average loneliness score that was calculated across all ill-health conditions that had at least 50 people in the group. We decided on 50 people as a minimum in a largely arbitrary manner, but one that was in-keeping with recommendations in psychology to have at least 50 items per cell of a design (Simmons et al., 2018). In addition, we also decided to rank order all ill-health conditions separately as a function of the condition-average loneliness score and then visually inspect where stroke “places” in the context of a wider set of ill-health conditions.

To compare loneliness scores following stroke to an average of all other ill-health conditions, we first calculated the mean average of all other ill-health conditions (n = 5550, x = 2.04, [1.99, 2.09]). We then ran an independent samples t-test, which showed that there was a small difference in the expected direction such that loneliness ratings were higher in the stroke group than the average ill-health condition group t(80.50) = 1.39, p = 0.08 difference in rating = 0.29, [-0.06, Inf], demonstrating a small effect and overlap between groups, d = 0.16, [-0.06, 0.38].

Further, in comparison with the other additional health conditions, history of stroke was demonstrated to be ranked 6^th^ out of 31 conditions in reported loneliness scores. Interestingly, the health conditions above stroke were, with the exception of stomach ulcer, all broadly psychological/neurological in nature (see Figure 2A). Further, there was one additional standout result - loneliness was particularly large in anxiety / depression compared to all other ill-health conditions, which is consistent with the well-established link between loneliness, anxiety and depression (Leary, 1990).

**Figure 2.**
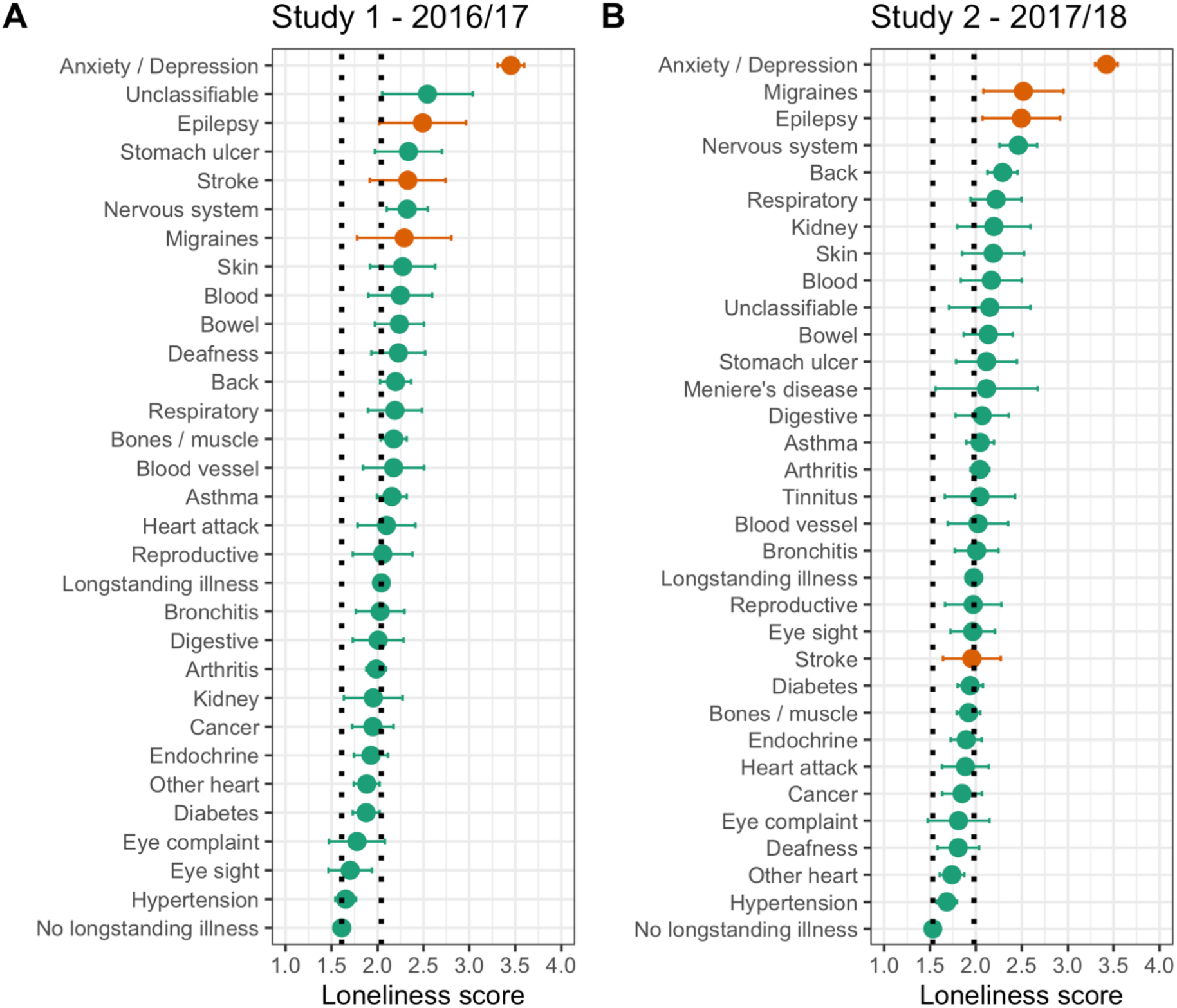
Forest plots depicting mean loneliness scores across a range of health conditions. Forest plots depicting mean loneliness scores and 95%CI for each health condition. (A) Results for Study 1 2016/17, (B) Results for Study 2 2017/18. Colour separates broadly defined brain (orange) and non-brain conditions (green). Dashed black lines demonstrate average loneliness score for healthy (left) and any longstanding illness group (right).

### Analysis of research question 2: Does CVA predict loneliness scores above demographic and social isolation variables

To address the second primary research question, a multivariable binary logistical regression analysis was completed in three stages. Model 1 included demographic variables such as age, gender, marital status and level of deprivation. Model 2 included variables relating to objective social isolation such as the number of people within home, internet access and population density. Finally, Model 3 included health condition status such as the presence of stroke, arthritis and any ill-health condition (‘ill’). Table 4 summarises the key findings from these models and the parameter estimates from the final model (model 3) are visualised in Figure 3.

**Table 3:** Between group differences in loneliness scores across Studies 1 and 2.

| *study* | *year* | *contrast* | *group1* | *group2* | *difference* | *conf.low* | *conf.high* | *t* | *df* | *p* | *d* | *d_conf.low* | *d_conf.high* |
| --- | --- | --- | --- | --- | --- | --- | --- | --- | --- | --- | --- | --- | --- |
| study1 | 16/17 | stroke > no longstanding illness | 2.33 | 1.61 | 0.72 | 0.37 | Inf | 3.44 | 80.10 | 0.00 | 0.45 | 0.23 | 0.67 |
| study1 | 16/17 | stroke > arthritis | 2.33 | 1.98 | 0.34 | -0.01 | Inf | 1.61 | 89.06 | 0.06 | 0.19 | -0.04 | 0.42 |
| study1 | 16/17 | stroke > ill | 2.33 | 2.04 | 0.29 | -0.06 | Inf | 1.39 | 80.50 | 0.08 | 0.16 | -0.06 | 0.38 |
| study1 | 16/17 | arthritis > no longstanding illness | 1.98 | 1.61 | 0.37 | 0.28 | Inf | 6.29 | 1545.78 | 0.00 | 0.23 | 0.16 | 0.30 |
| study2 | 17/18 | stroke > no longstanding illness | 1.96 | 1.53 | 0.43 | 0.16 | Inf | 2.66 | 120.56 | 0.00 | 0.27 | 0.09 | 0.46 |
| study2 | 17/18 | stroke > arthritis | 1.96 | 2.04 | -0.09 | -0.36 | Inf | -0.53 | 141.81 | 0.70 | -0.05 | -0.24 | 0.14 |
| study2 | 17/18 | stroke > ill<sup>†</sup> | 1.96 | 1.98 | -0.02 | -0.29 | Inf | -0.12 | 121.69 | 0.55 | -0.01 | -0.19 | 0.17 |
| study2 | 17/18 | arthritis > no longstanding illness | 2.04 | 1.53 | 0.51 | 0.42 | Inf | 9.13 | 1757.69 | 0.00 | 0.32 | 0.26 | 0.38 |

<sup>†</sup>ill = any longstanding illness. Conf.low = lower bound of 95% confidence interval. Conf.high = upper bound of 95% confidence interval. t = t statistic, df = degrees of freedom, p = p value, d = Cohen's d.

**Table 4:**
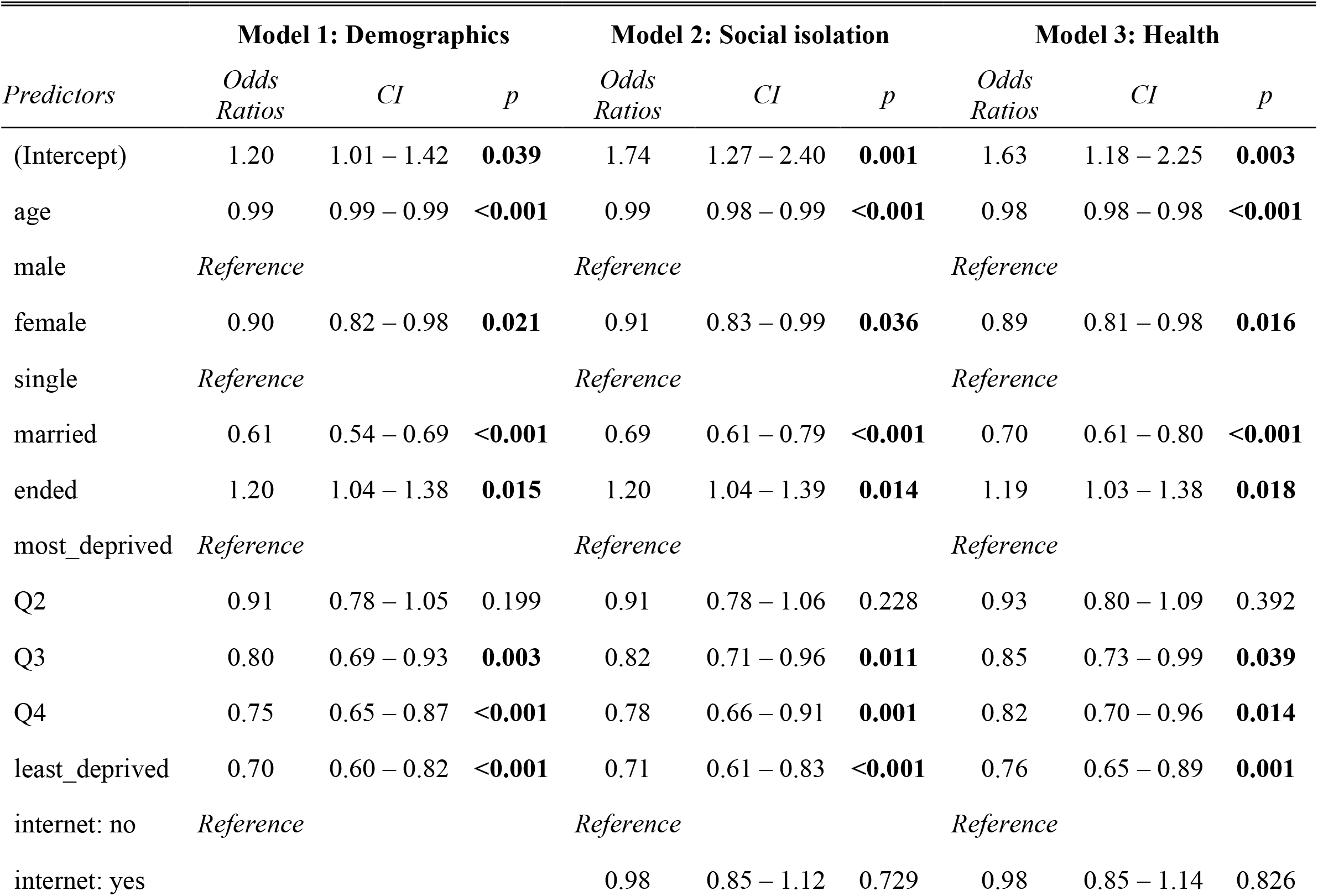
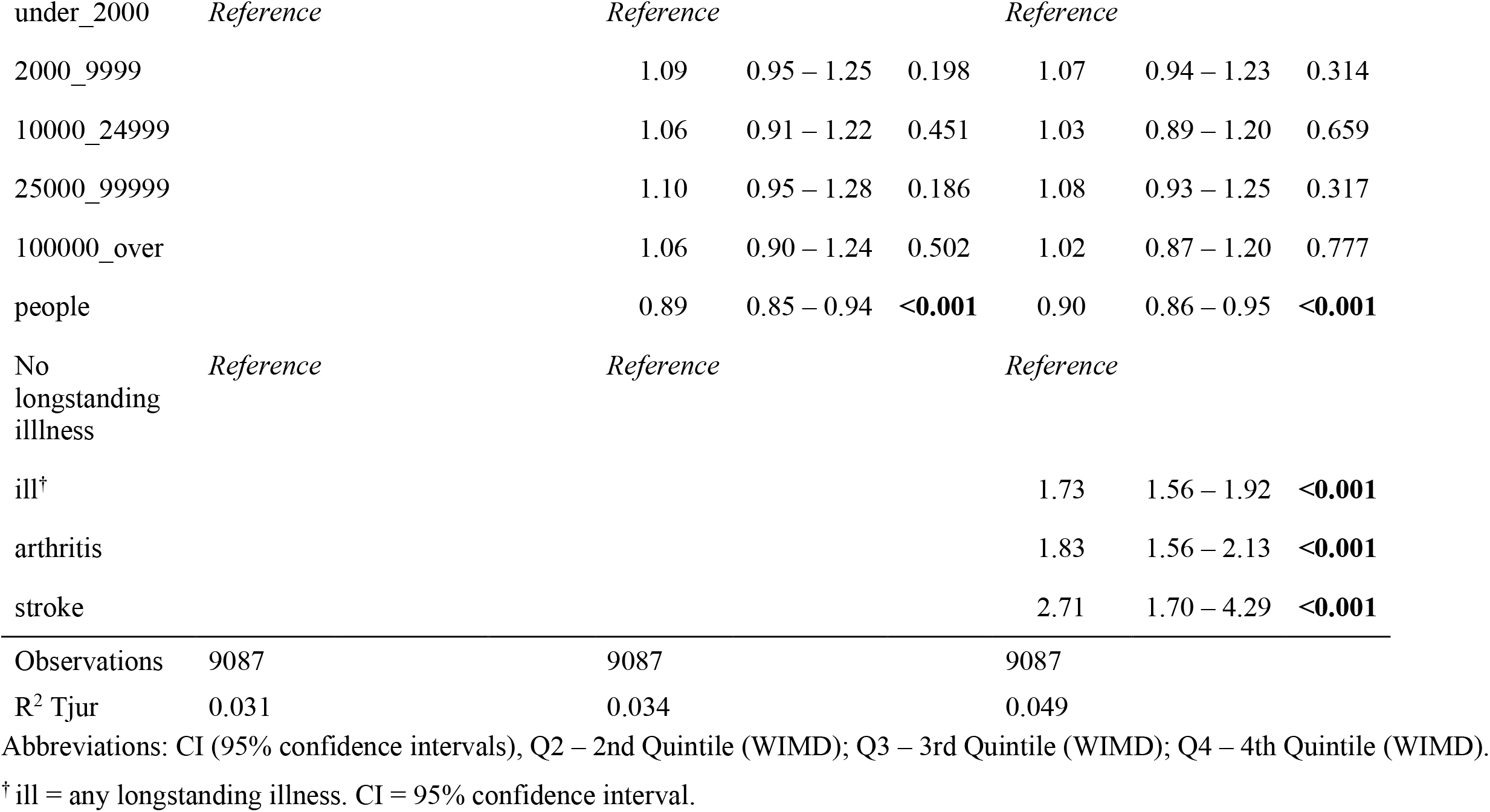
Study 1 logistic regression summary data.

**Figure 3.**
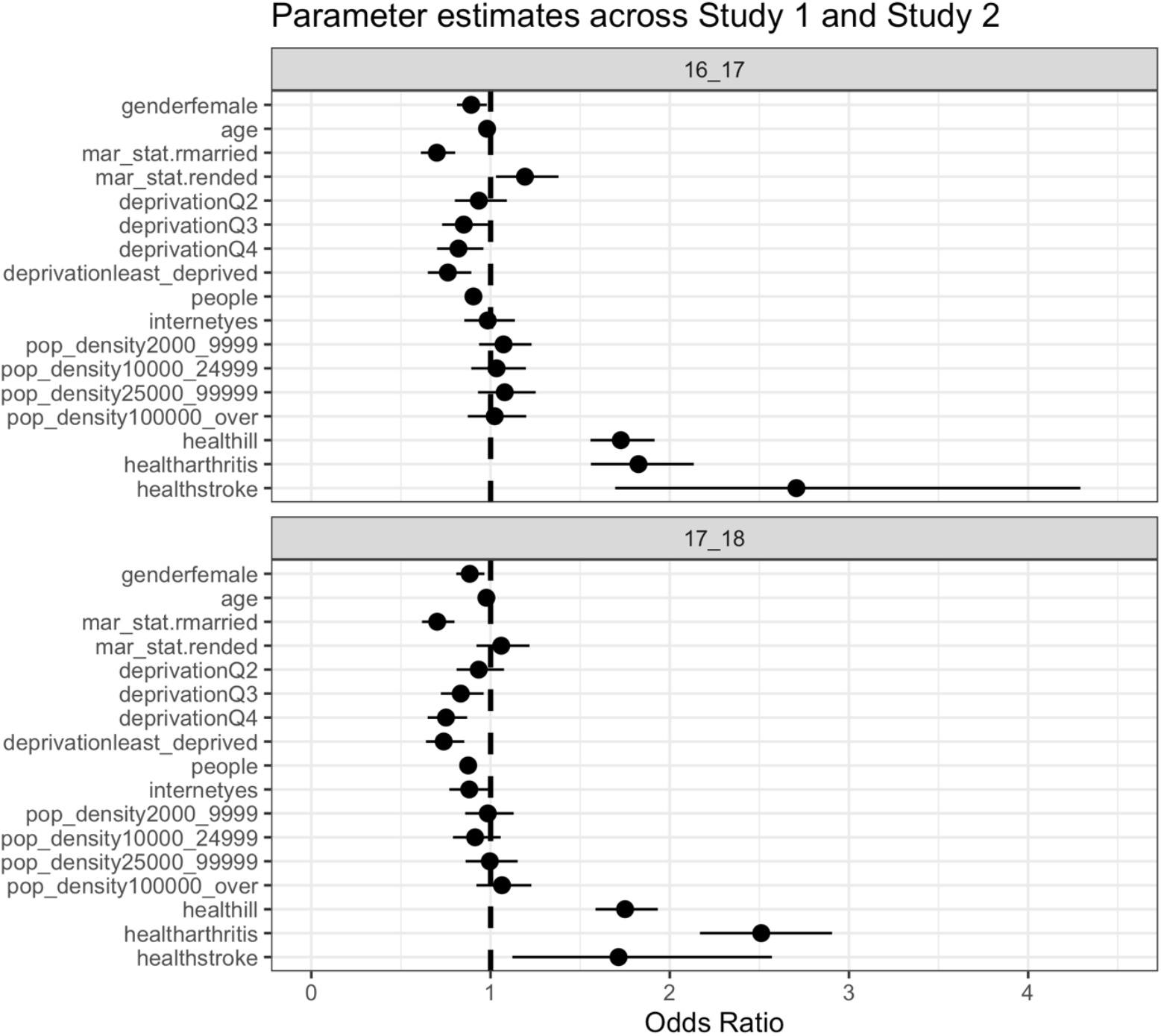
Odds ratios for predictors of interest across Studies 1 and 2. Parameter estimates (odds ratios) across Studies 1 (16_17) and 2 (17_18). Odds ratios were derived using logistic regression with self-reported loneliness as the dependent variable. The loneliness scale was scored from 0-6, which was recoded as zero if less than three and one otherwise. The reference category for gender was male. Age was included as a continuous variable. Mar_stat = marital status, which included three levels: single (reference category), married and ended. There were 5 levels of deprivation, with the most deprived group included as the reference category (Q = quintiles). People refers to the number of people within the household and was included as a continuous variable. Internetyes refers to those individuals who reported having an internet connection (reference category = no connection). Pop_density refers to the population density of the household’s location with the sparsest density (<2000 people) as the reference category. Health refers to health status/condition with the reference category being healthy individuals who report no longstanding illness. Point estimates (dots) represents the mean estimate odds ratios and whiskers represent 95% confidence intervals. For more information see Tables 4 and 5.

Model 1, which included all demographic variables, explained a small amount of variance in loneliness scores (Tjur R^2^ 0.031). Categories of marital status such as being ‘separated’, ‘divorced’ or ‘widowed’ were associated with higher reported loneliness (OR = 1.20, [1.04, 1.38]). In contrast, being married (OR = .61, [.54, .69]) was associated with lower reported loneliness. Similarly, effects in this direction were demonstrated for other demographic variables such as gender (female [OR = 0.90, [.82, .98]), age (OR = .99, [.99, .99)] and across the three least deprived areas; Quintile 3 (OR = .80, [.69, .93]), Quintile 4 (OR = .75, [.65, .87]) and Quintile 5 (OR = .70, [CI .60 - .82]).

**Table 5:**
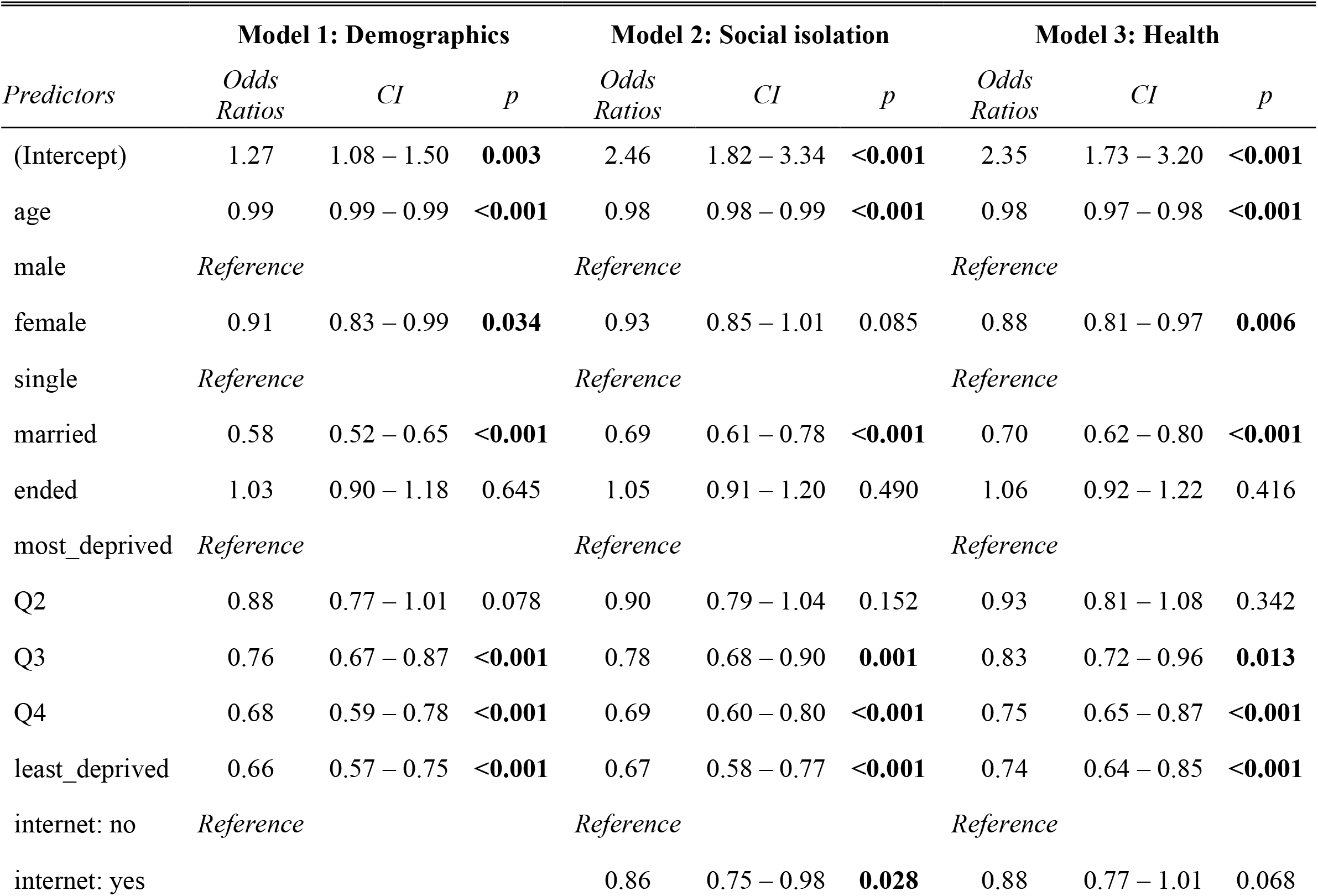
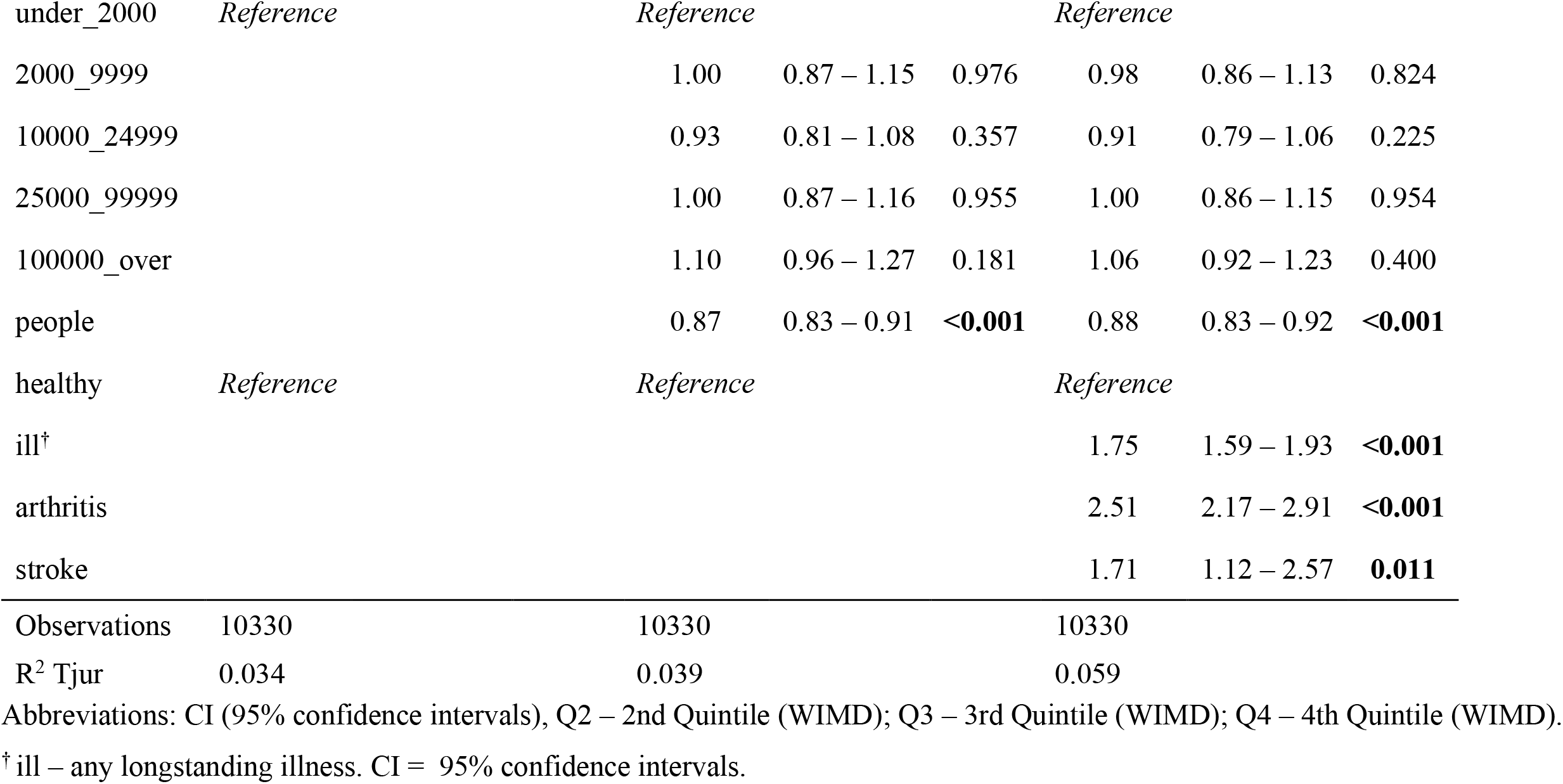
Study 2 logistic regression summary data.

Model 2, which included variables relating to social isolation, also explained a small amount of the overall variance in loneliness scores (Tjur R^2^ 0.034). One variable that predicted loneliness in model 2 was the number of people in the household, with greater numbers associated with reduced reported loneliness (OR = .89, [.85, .94]). Other factors such as population density and access to internet connection were found to have a negligible impact on loneliness (see Table 4 for details).

The inclusion of health status (stroke, arthritis, & any ill-health condition) in Model 3 increased the explanatory power of the overall model (Tjur R^2^ 0.049). Stroke was found to be the largest predictor of reported loneliness across health conditions such that it substantially increased reported loneliness over two-fold (OR = 2.71, [1.70, 4.29]). Multicolinearity amongst the predictor variables was generally low, with variance inflation factors calculated using the ‘vif’ function in the R package ‘car’ all under 3, but with most under 1.5. Further details can be found with the analysis scripts hosted on the open science framework.

### Confirmatory phase (Study 2)

Based on the analyses performed in Study 1, we refined our analytical strategy to focus on providing confirmatory evidence for our key hypotheses of interest. Like Study 1, we pre-registered our analysis questions, hypotheses and key analytical approach (https://aspredicted.org/xq5bd.pdf). In general, we largely used the same analytical approach to Study 1, but we benefited from being in a more informed position to test predictions based on the evidence emerging from Study 1.

### Describing loneliness across our groups of interest

The total prevalence of loneliness reported, as defined by scores of 3 or over on the DJGLS, was 30% in the stroke group, 37.6% in the arthritis group and 25% in the no longstanding illness group. In addition, reported loneliness ratings across the three groups were as follows: stroke (n = 117, x = 1.96, [1.75, 2.71]); arthritis (N = 1408, x = 2.04, [1.94, 2.14]), and; no longstanding illness(n = 5347, x = 1.53, [1.49, 1.57]) (Table 2, Figure 4).

**Figure 4.**
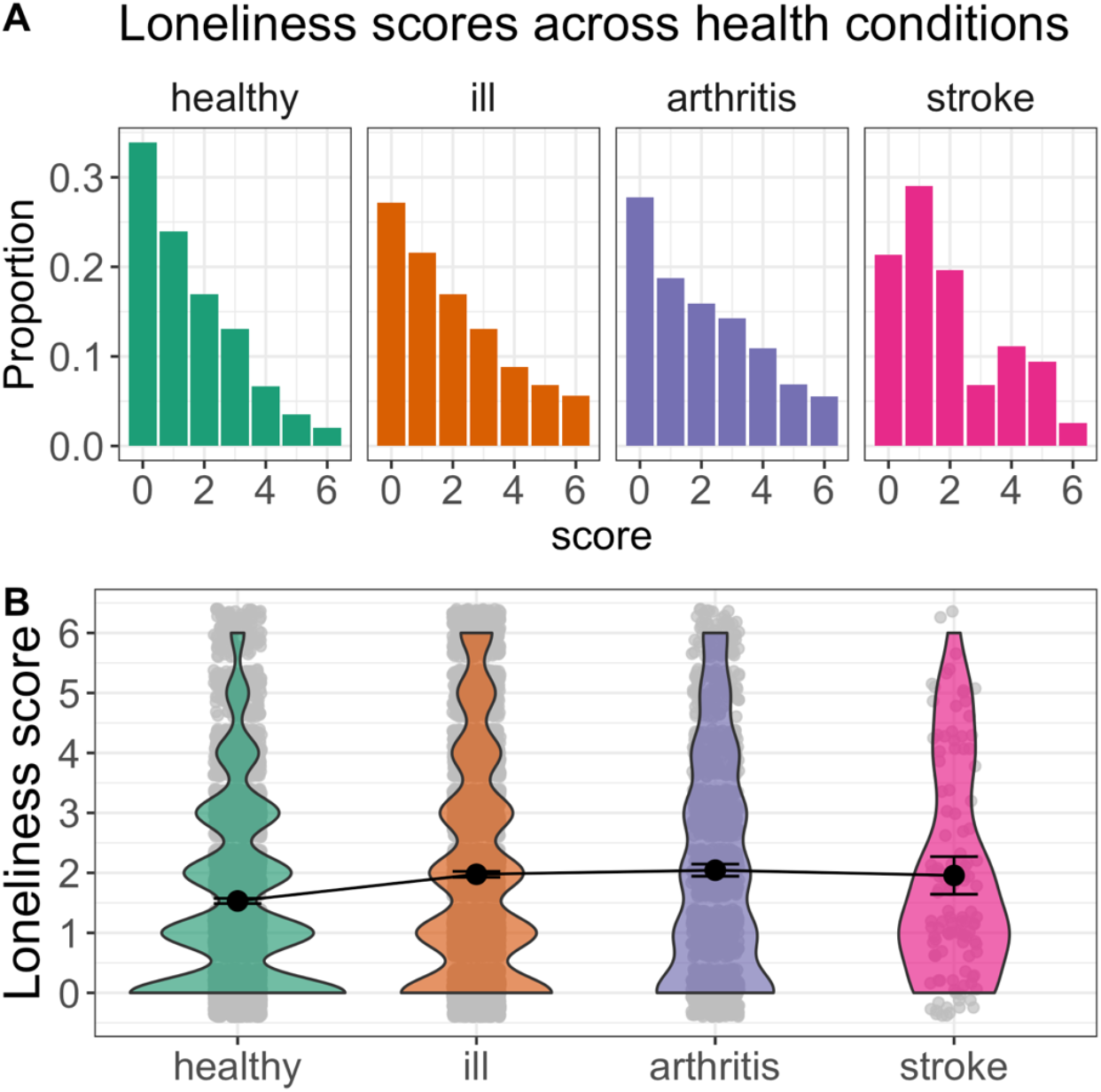
Loneliness scores across health conditions in Study 2. Histograms (A) and violin plots (B) of loneliness scores in Study 2. A) Histogram representing the proportion of reported loneliness score for each health condition. B) Violin plot representing the mean and 95%CI for each health control group in addition to the distribution of scores. Healthy = no longstanding illness, Ill = any longstanding illness.

### Analysis of research question 1: To what extent do loneliness ratings differ between health conditions

Three independent samples t-tests were performed to estimate pair-wise group differences of interest. First, we compared loneliness scores between CVA and NLI groups. The stroke group reporting higher loneliness than the no longstanding illness group t(120.56) = 2.66, p < 0.00, difference in rating = 0.43, [0.16, Inf], demonstrating a small effect with no overlap between groups, d = 0.27, [0.09, 0.46]

Second, we compared the stroke group to the arthritis group. In contrast to Study 1, the stroke group did not show a clear difference in loneliness to the arthritis group t(141.81) = 0.53, p = 0.70, difference in rating = -0.09, [-0.36, Inf], demonstrating no effect with large overlap between group, d = 0.05, [-0.24, 0.14].

Third, we compared the arthritis group to the no longstanding illness group. The arthritis group again reported higher loneliness levels compared to the no longstanding illness group t(1757.69) = 9.13, p = 0.00, difference in rating = 0.51, demonstrating a small effect with no overlap between groups, d = 0.32, [0.26, 0.38].

Like Study 1, we also compared loneliness ratings to a wider set of ill-health groups. First, we compared loneliness ratings in the stroke group to an average loneliness score that was calculated across all ill-health conditions that had at least 50 people in the group. In addition, we also ranked all ill-health conditions separately as a function of the condition-average loneliness score and then visually inspect where stroke “places” in the context of a wider set of ill-health conditions.

To compare loneliness scores following stroke to an average of all other ill-health conditions, we first calculated the mean average of all other ill-health conditions (N = 5971, x = 1.98, [1.93, 2.03]). We then ran an independent samples t-test, which did not show clear evidence for a difference in loneliness ratings between the stroke group compared to the average ill-health condition t(121.69) = 0.12, p = 0.55, difference in rating = -0.02, demonstrating no small effect and large overlap between group, d = 0.01, [-0.19, 0.17].

When we rank ordered the average loneliness score per health condition, several results became immediately obvious. First, neurological conditions in general tended to cluster near the top of the list and loneliness was particularly high in anxiety / depression compared to all other ill-health conditions (Leary, 1990). The pattern of these results closely replicate Study 1 and thus provide reasonably strong confirmatory evidence. Second, and in contrast to Study 1, the stroke group was positioned in the middle of the pack and overlapped almost perfectly with the average rating for all ill-health conditions (Figure 2B). Therefore, Study 2 does not confirm the position that stroke results in demonstrably higher levels of loneliness than a range of other ill-health conditions.

### Analysis of research question 2: Does stroke predict loneliness scores above demographic and social isolation variables

To address our second research question in Study 2, we used the same multivariable binary logistical regression analysis. We calculated three models. Model 1 included demographic variables, Model 2 included social isolation variables and finally Model 3 included presence of stroke, arthritis and also any longstanding health condition (‘ill’). Table 5. summarises the key findings from these models and the parameter estimates from the final model (model 3) are visualised in Figure 3.

Model 1 was found to explain a small amount of the variance in reported loneliness (R^2^ Tjur 0.034). In line with the exploratory phase findings, being married (OR = .58, [.52, .65]) and being younger in age (OR = .99, [.99, .99]) were found to reduce likelihood of reported loneliness. The replication of the exploratory results was also demonstrated when measuring the effect ‘level of deprivation’; Quintile 3 (OR = .76, [.67, .87]), Quintile 4 (OR = .68, [.59, .78]) and Quintile 5 (OR = .66, [.57, .75]). These results confirm that lower levels of deprivation are associated with a reduced likelihood of reported loneliness.

Model 2, which additionally included social isolation variables, also explained a small amount of the variance in reported loneliness (R^2^ Tjur 0.039). In line with findings from Study 1, the number of people in household (OR = .87, [.83, .91]) was found to be a significant predictor of loneliness, with more people in the house reducing the likelihood of reporting loneliness. In contrast to the exploratory phase, having internet access decreased the likelihood of reporting loneliness (access to internet [OR = .86, [.75, .98]).

The introduction of health status in Model 3 increased the explanatory power of the model (R^2^ Tjur 0.059). In addition, the presence of stroke was found to be a significant predictor of reported loneliness (OR = 1.71, [1.12, 2.57]). However, in contrast to Study 1, the presence of arthritis appeared to be the biggest predictor of reported loneliness across the health conditions included in the model (OR = 2.51, [2.17, 2.91]). Multicolinearity amongst the predictor variables was generally low, with variance inflation factors calculated from the using the ‘vif’ function in the R package ‘car’ all under 3, but with most under 1.5. Further details can be found with the analysis scripts hosted on the open science framework.

### Meta-analysis

A meta-analysis was completed to provide a weighted average of the key group differences across Studies 1 and 2. We estimated the weighted average in original units, which means points on the loneliness scale that ranges from 0-6. An overall mean difference of 0.54 [0.26 – 0.82] was found between those with stroke and those with no longstanding illnesses (Figure 5A). The direction of the difference is consistent across both studies such that the stroke group report numerically higher levels of loneliness than the no longstanding illnesses group.

**Figure 5.**
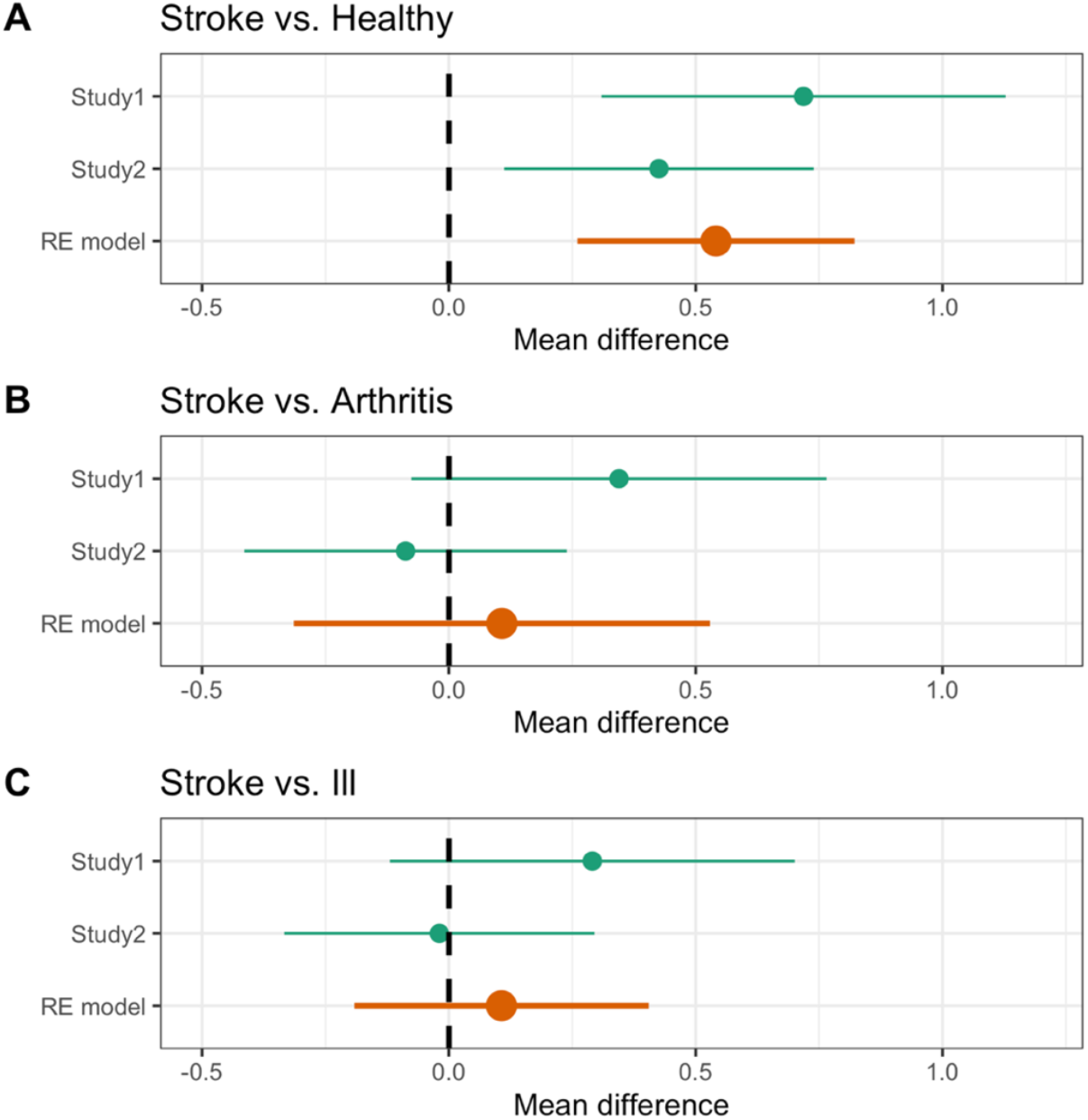
Meta-analytical results for between group comparisons. Meta-analytical results. A) Mean difference in loneliness scores between those with a stroke and those with no longstanding illnesses. B) Mean difference in loneliness scores between those with a stroke and those with Arthritis. C) Mean difference in loneliness scores between those with a stroke and those with any longstanding illness. Orange indicates the random effects (RE) model estimates for mean difference scores across both studies (Study 1 and 2). Healthy = no longstanding illness, Ill = any longstanding illness.

In contrast, a negligible effect was demonstrated between stroke and arthritis (mean difference = 0.11 [-0.31 – 0.53]; Figure 5B) and between stroke and those reporting any longstanding illness (mean difference = 0.11[-0.19 – 0.40]; Figure 5C). Although the effects are in the predicted direction, the interval estimates show considerable overlap with zero and the difference is too small to have a reasonable level of confidence regarding the effects.

## Discussion

This is the first large-scale study to estimate levels of loneliness within the stroke population, whilst accounting for objective measures of social isolation. Across both datasets, the results consistently demonstrate that loneliness is elevated following stroke compared to healthy individuals. Moreover, the impact of having a stroke is not accounted by objective measures of social isolation such as marital status or the number of people in a household. Therefore, these results extend prior work that showed stroke is associated with objective measures of social isolation (Valtorta Kanaan, Gilbody, Ronzi, et al., 2016), by showing how the impact of having a stroke is also tied to the interpretation of your social environment. This finding is important because of the substantial impact that loneliness specifically, rather than objective measures of social isolation, has on increases in morbidity and mortality (Holt-Lunstad et al., 2010; 2015).

To help contextualise our main findings, we compare the impact of stroke to other predictors of loneliness, as well as to other illnesses. Across both studies, stroke survivors were at least 70% more likely to report experiencing higher levels of loneliness compared to healthy individuals. The size of such results is comparable to the impact of other factors that influence loneliness, such as marital status, where being married was related to a reduction in loneliness of approximately 30%. Such comparisons mean that brain injury may have a non-trivial impact on loneliness, which is similar or larger in magnitude than other factors that impact levels of loneliness. As such, we do not think it can be ignored or downplayed as a statistical irrelevance; instead, it may have a meaningful impact on quality of life.

By contrast, the relationship between stroke and loneliness was not consistently greater than the strength of the relationship between loneliness and other comparison illnesses, such as arthritis or an average of all illnesses. On current evidence, therefore, we cannot conclude that loneliness levels are elevated post-stoke more than following a range of other illnesses. It is worth noting, however, that our exploratory analysis of wider illnesses shows that across both datasets, those with loosely-defined brain-related illnesses (e.g., depression/anxiety, epilepsy, migraines) consistently reported experiencing the highest levels of loneliness. Indeed, out of approximately 30 illnesses, brain-related illnesses were in the top 5 each time (see Figure 2). This may not be surprising given the pathology occurs in the organ that is responsible for social behaviour. However, further research is needed to explore this tentative finding by comparing across different illness groups in a more comprehensive manner.

The prevalence rates established in the current study suggest that 30% - 44% of those with a history of stroke report to be lonely. This is a three-fold increase compared to findings from similar studies examining loneliness in a typical/healthy population (Beurel et al 2017). Similarly, it is a two-fold increase in comparison to recent studies examining loneliness in general primary care outpatients (Mullen, Tong, Sabo et al. 2019). From a clinical perspective, therefore, these findings suggest that at least one in three stroke survivors that clinicians encounter may be suffering from loneliness. To further emphasise the newly found prevalence of loneliness in this population; it is comparable to, even exceeding, the prevalence rates of other psychological/emotional sequalae such as depression (30%) and anxiety (25%) (Campbell, Burton, Murray, Holmes et al 2013; Paolucci, 2008), which are routinely screened for in clinical assessment.

Of course, depression, anxiety and loneliness are not mutually exclusive and are likely interrelated (Matthews, Danese, Wertz, 2016), as broadly demonstrated in our results section. Future research that unpicks these relationships would be valuable. Irrespective of the likely complex relationship between loneliness and other variables, however, it is clear that if clinicians ignore the social consequences that follow from a stroke, it is likely to have serious health implications. Further, the results provide empirical support for the idea that novel clinical solutions such as social prescriptions need to be embedded into clinical practice alongside standard treatment. Importantly, our results also suggest that such social prescriptions should target developing a greater quality, rather than quantity, of social connections (Bruine de Bruin, Parker, & Strough, 2020; Cacioppo & Patrick, 2008; Hawkley et al., 2008).

### Strengths, limitations and future directions

The use of multi-cohort survey data from a large representative sample of an entire country is a significant a strength of the current study. To date, there has not been a large-scale study examining loneliness in the stroke population. By using open science principles, such as pre-registration and replication, as well as using larger samples sizes (Munafo et al., 2017; Open Science Collaboration, 2015; Simmons et al., 2018; Zwaan et al., 2018), we have provided the most rigorous evidence to date that addresses this research question. In addition, by making the analysis pipeline available for others to use, we hope to facilitate a move towards a more cumulative science by enabling others to build upon our work in meta-analyses and to power future studies.

We also acknowledge several limitations from using survey data. Like all survey designs, the data gained from the participants relating to health condition and loneliness levels were self-reported and could be liable to response bias. If individuals responded in misleading or untruthful ways in order to conform with social expectations, it may result in an underestimation or overestimation of our effects. In addition, the use of secondary data means that the authors were not involved in the design of the survey. Therefore, potentially important factors such as the topography of the stroke (e.g., time since stroke, as well as location and severity of stroke) were not measured and their potential impact on loneliness is therefore unknown. As such, future studies may examine whether the nature of the stroke influences loneliness outcomes. Preliminary evidence suggests that the time since the occurrence of a stroke may be important in the evolution of loneliness such that loneliness may become more prevalent in the later stages of recovery (Harrick et al., 1994). The location of the stroke may also prove informative when examining for any cognitive correlates involved in loneliness. Social behaviour requires an integration of many, if not all, cognitive processes. Therefore, disturbances to distinct neuroanatomical areas may shine a light on the crucial cognitive underpinnings associated with loneliness.

A final limitation of the survey design is that it cannot be used to infer causation. As discussed previously, studies have demonstrated that loneliness has been associated with increased mortality and morbidity (Holt-Lunstad et al., 2010; 2015). A recent meta-analysis demonstrated that poor social relationships were associated with a 32% increased risk (relative risk) of stroke (Valtorta, Kanaan, Gilbody, Ronzi, et al., 2016). Taken together, it is difficult to establish causation, as it may be that the loneliness preceded stoke or that loneliness and stroke are mutually reinforcing. Longitudinal studies may help to establish the temporal relationship between loneliness and stroke in the stroke population.

## Conclusion

This study demonstrates that the perception that your social needs are not being met - loneliness - is elevated in the stroke population. Our results indicate that one in three people with a history of stroke report to be lonely, which emphasises the need for more public and clinical awareness of loneliness in those with history of stroke. As with other emotional/psychological conditions such as anxiety and depression, the findings suggest that loneliness should be screened for by clinicians working with this clinical population. This should be considered crucial from a neurorehabilitation perspective given that loneliness has been demonstrated to mediate the trajectory of recovery, and deterioration, in other diseases and health conditions (Friedler et al 2015; Grande et al 2018).

## Data Availability

This project involves secondary data analysis and the data are available from the UK Data Service.

https://ukdataservice.ac.uk

## Acknowledgments

This research was performed as part of an all-Wales Economic and Social Research Council (ESRC) Doctoral Training Centre PhD studentship (awarded to RR and RC, PhD student: CB).

## References

1. Beutel, M. E., Klein, E. M., Brähler, E., Reiner, I., Jünger, C., Michal, M., … Tibubos, A. N. (2017). Loneliness in the general population: prevalence, determinants and relations to mental health. BMC psychiatry, 17(1), 97. Doi:10.1186/s12888-017-1262-x

2. Brittain, K., Kingston, A., Davies, K., Collerton, J., Robinson, L., Kirkwood, T., Bond, J. Jagger, C. (2017). An Investigation Into The Patterns Of Loneliness And Loss In The Oldest Old – Newcastle 85 Study. Ageing And Society, 37(1), 39–62. Doi:10.1017/S0144686x15001142

3. Bruine de Bruin, W., Parker, A. M., & Strough, J. (2020). Age differences in reported social networks and well-being. Psychology and Aging, 35(2), 159–168. doi:10.1037/pag0000415

4. Cacioppo, J. T., & Patrick, W. (2008). Loneliness: Human nature and the need for social connection. W W Norton & Co

5. Cacioppo, J. T., & Hawkley, L. C. (2009). Perceived Social Isolation and Cognition. Trends in Cognitive Sciences, 13, 447–454.

6. Cacioppo, J. T., Cacioppo, S., Capitanio, J. P., & Cole, S. W. (2015). The Neuroendocrinology of Social Isolation. Annual Review of Psychology, 66(1), 733–767. doi:doi:10.1146/annurev-psych-010814-015240

7. Cacioppo, S., Grippo, A. J., London, S., Goossens, L., & Cacioppo, J. T. (2015). Loneliness: clinical import and interventions. Perspectives on psychological science: a journal of the Association for Psychological Science, 10(2), 238–249. https://doi.org/10.1177/1745691615570616

8. Campbell Burton C. A., Murray J., Holmes J., Astin F., Greenwood D., Knapp P. (2013) Frequency of anxiety after stroke: a systematic review and meta-analysis of observational studies. International Journal of Stroke. 8(7):545–559.

9. Cumming, G. (2012). Understanding the new statistics: Effect sizes, confidence intervals, and meta-analysis. New York: Routledge

10. Cumming, G. (2014). The New Statistics: Why and How. Psychological Science, 25(1), 7–29.

11. Friedler, B., Crapser, J., & McCullough, L. (2015). One is the deadliest number: the detrimental effects of social isolation on cerebrovascular diseases and cognition. Acta neuropathologica, 129(4), 493–509. https://doi.org/10.1007/s00401-014-1377-9

12. Gierveld, J. D. J, Tilburg, T. V. (2006). A 6-Item Scale for Overall, Emotional, and Social Loneliness: Confirmatory Tests on Survey Data. Research on Aging, 28, 582–598.

13. Gigerenzer, G. (2018). Statistical Rituals: The Replication Delusion and How We Got There. Advances in Methods and Practices in Psychological Science, 1(2), 198–218

14. Grande, G., Vetrano, D. L., Cova, I., Pomati, S., Mattavelli, D., Maggiore, L., … Rizzuto, D. (2018). Living Alone and Dementia Incidence: A Clinical-Based Study in People With Mild Cognitive Impairment. Journal of Geriatric Psychiatry and Neurology, 31(3), 107–113.

15. Helme, M & Brown, Z (2018). National Survey for Wales 2017-18 Technical Report. Welsh Gvernment. Retrieved from https://gov.wales/sites/default/files/statistics-and-research/2019-05/national-survey-for-wales-technical-report-2017-18_0.pdf

16. Hall, E.T. (1963) A System for the Notation of Proxemic Behavior. American Anthropologist. 65:1003–1026.

17. Harrick, L., Krefting, L. Johnston, J., Carlson, P., & Minnes, P. (1994) Stability of functional outcomes following transitional living programme participation: 3-year follow-up, Brain Injury, 8:5, 439–447.

18. Hawkley, L. C., Hughes, M. E., Waite, L. J., Masi, C. M., Thisted, R. A., & Cacioppo, J. T. (2008). From social structural factors to perceptions of relationship quality and loneliness: The Chicago Health, Aging, and Social Relations Study. Journals of Gerontology, Series B: Psychological Sciences and Social Sciences, 63, S375–S384

19. Hienrich, L. M., & Gullone, E. (2006). The clinical significance of loneliness: a literature review. Clin Psychol Rev. 26(6), 695–718.

20. Holt-Lunstad J, Smith TB, Layton JB (2010) Social Relationships and Mortality Risk: A Meta-analytic Review. PLoS Med 7(7): e1000316. https://doi.org/10.1371/journal.pmed.1000316

21. Holt-Lunstad, J., Smith, T. B., Baker, M., Harris, T., & Stephenson, D. (2015). Loneliness andsocial isolation as risk factors for mortality: A meta-analytic review. Perspectives on Psychological Science, 10, 227–237

22. Holwerda T. J., Deeg D. J. H., Beekman A. T. F. ….Schoevers, R. A. (2014). Feelings of loneliness, but not social isolation, predict dementia onset: results from the Amsterdam Study of the Elderly (AMSTEL) Journal of Neurology, Neurosurgery & Psychiatry, 85, 135–142.

23. Kane, M. & Cook, L (2013). Dementia 2013: The hidden voice of loneliness. Alzheimer’s Society.

24. Kruschke, J.K., & Liddell, T.M. (2018). The Bayesian New Statistics: Hypothesis testing, estimation, meta-analysis, and power analysis from a Bayesian perspective. Psychon Bull Rev 25, 178–206. https://doi.org/10.3758/s13423-016-1221-4

25. Leary, M. R. (1990). Responses to social exclusion: Social anxiety, jealousy, loneliness, depression, and low self-esteem. Journal of Social and Clinical Psychology, 9(2), 221–229.

26. Matthews, T., Danese, A., Wertz, J., Odgers, C. L., Ambler, A., Moffitt, T. E., & Arseneault, L. (2016). Social isolation, loneliness and depression in young adulthood: a behavioural genetic analysis. Social psychiatry and psychiatric epidemiology, 51(3), 339–348. doi:10.1007/s00127-016-1178-7

27. McElreath, R. (2016). Statistical Rethinking: A Bayesian Course with Examples in R and Stan CRC Press

28. Mullen, R. A., Tong, S. T, Sabo, R.T et al. (2019). Loneliness in primary care patients: a prevalence study. Ann Fam Med. 17, 108–115

29. Munafò, M., Nosek, B., Bishop, D. et al.(2017). A manifesto for reproducible science. Nat Hum Behav 1, 0021.

30. Nelson, L. D., Simmons, J., Simonsohn, U. (2018). Psychology’s Renaissance Annual Review of Psychology 2018 69:1, 511–534

31. Open Science Collaboration (2015) Estimating the reproducibility science. Science, 349; 6251

32. Paolucci S. (2008). Epidemiology and treatment of post-stroke depression. Neuropsychiatric disease and treatment, 4(1), 145–154. doi:10.2147/ndt.s2017

33. Perissinotto, C. M., Cenzer, I. S., & Covinsky, K. E. (2012). Loneliness in older persons: a predictor of functional decline and death. Archives of internal medicine, 172(14), 1078–1084.

34. Perissinotto, C. M., & Covinsky, K. E. (2014). Living alone, socially isolated or lonely— What are we measuring? Journal of General Internal Medicine, 11, 1429–31.

35. Qualter, P., Vanhalst, J., Harris, R., van Roekel, E., Lodder, G., Bangee, M., … Verhagen, M. (2015). Loneliness across the life span. Perspectives on Psychological Science, 10(2), 250–264

36. Rafnsson S. B., Orrell, M,. D’Orsi, E., Hogervorst, E., & Steptoe, A. (2017). Loneliness, social integration, and incident dementia over 6 years: Prospective findings from the English Longitudinal Study of Ageing. J Gerontol B Psychol Sci Soc. 75, 114–124.

37. Simmons, J. P., Nelson, L. D., & Simonsohn, U. (2011). False-positive psychology: Undisclosed flexibility in data collection and analysis allows presenting anything as significant. Psychological Science, 22, 1359–1366

38. Simmons, J. P., Nelson, L. D., & Simonsohn, U. (2018). False-Positive Citations. Perspectives on Psychological Science 13(2), 255–259. https://doi.org/10.1177/1745691617698146

39. Stroke Association (2018) State of The Nation: Stroke statistics. Accessed 18th June 2020 from https://www.stroke.org.uk/sites/default/files/state_of_the_nation_2018.pdf

40. Valtorta, N. K., Kanaan, M., Gilbody, S., Ronzi, S., & Hanratty, B. (2016). Loneliness and social isolation as risk factors for coronary heart disease and stroke: systematic review and meta-analysis of longitudinal observational. Heart, 102, 1009-16. studies.

41. Valtorta, N. K., Kanaan, M., Gilbody, S., & Hanratty, B. (2016). Loneliness, social isolation and social relationships: what are we measuring? A novel framework for classifying and comparing tools. BMJ open, 6(4), e010799.

42. van Vollenhoven R. F. (2009). Sex differences in rheumatoid arthritis: more than meets the eye… BMC medicine, 7, 12. doi:10.1186/1741-7015-7-12

43. Viechtbauer, W. (2010). Conducting meta-analyses in R with the metafor package. Journal of Statistical Software, 36(3), 1–48. https://www.jstatsoft.org/v036/i03.

44. Zwaan, R., Etz, A., Lucas, R., & Donnellan, M. (2018). Making replication mainstream. Behavioral and Brain Sciences, 41, E120. doi:10.1017/S0140525x17001972

